# Early phases of COVID-19 are characterized by a reduction of lymphocyte populations and the presence of atypical monocytes

**DOI:** 10.1101/2020.05.01.20087080

**Authors:** Andrea Lombardi, Elena Trombetta, Alessandra Cattaneo, Valeria Castelli, Emanuele Palomba, Mario Tirone, Davide Mangioni, Giuseppe Lamorte, Maria Manunta, Daniele Prati, Ferruccio Ceriotti, Roberta Gualtierotti, Giorgio Costantino, Stefano Aliberti, Vittorio Scaravilli, Giacomo Grasselli, Andrea Gori, Laura Porretti, Alessandra Bandera, for the COVID-19 Network

## Abstract

**Background:** Severe acute respiratory syndrome coronavirus 2 is a recently discovered pathogen responsible of coronavirus disease 2019 (COVID-19). The immunological changes associated with this infection are largely unknown.

**Methods:** We evaluated the peripheral blood mononuclear cells profile of 63 patients with COVID-19 at diagnosis and the presence of association with inflammatory biomarkers and 28-days mortality.

**Results:** Lymphocytopenia was present in 51 of 63 (80.9%) patients. This reduction was mirrored also on CD8+ lymphocytes (128 cells/μL), natural killer cells (67 cells/μL) and natural killer T cells (31 cells/μL). Monocytes were preserved in total number but displayed a subpopulation composed mainly of cells with a reduced expression of both CD14 and HLA-DR. A direct correlation was found between serum values of IL-6 and the frequency of Th2 lymphocytes (R=0.17; *p*=0.04) but not with the monocytes count (R=0.01; *p*=0.60). Patients who died in the 28 days from admission (N=10, 15.9%), when compared to those who did not, displayed lower mean values of CD3+ (*p*=0.028) and CD4+ cells (*p*=0.042) and higher mean percentages of CD8+/CD38+/HLA-DR+ lymphocytes (*p*=0.026).

**Conclusions:** The early phases of COVID-19 are characterized by lymphocytopenia, predominance of Th2 lymphocytes and less immunocompetent monocytes, which include atypical mononuclear cells.

**eTOC:**

-At diagnosis patients with COVID-19 have lymphocytopenia
-Monocytes with both normal or altered scatter properties display a reduced expression of CD14 and HLA-DR in most of COVID-19 patients
-Patients who die in the 28 days from admission have lower values of CD3+ and CD4+ cells and higher percentages of activated CTL cells compared to those who survive

## Introduction

Severe acute respiratory syndrome coronavirus 2 (SARS-CoV-2) is a new beta coronavirus identified in China in December 2019 which is responsible of coronavirus disease 2019 (COVID-19).(World Health Organization (WHO), 2020) The virus rapidly spread worldwide, and it is responsible of a pandemic which is exerting a tremendous pressure on national health systems.(Villa et al., 2020)

Given the novelty of this virus, how the immune system deals with it is largely unknown. Preliminary studies from China highlighted a reduction of lymphocytes count, particularly of CD4+ T and CD8+ T cells, an increase of the neutrophils-to-lymphocytes ratio (NLR), a concomitant decrease in interferon gamma (IFN-*γ*) production and an association with elevated inflammatory markers.(Chuan et al., 2020; Chen et al., 2020; Liu et al., 2020) An excessive pro-inflammatory cytokines release profile was confirmed also by a transcriptomic study performed on peripheral blood mononuclear cells (PBMC).(Xiong et al., 2020) Also innate immunity cells are involved. Indeed, Zheng and colleagues demonstrated an upregulation of the inhibitory receptor NKG2A expression on natural killer cells (NK) and cytotoxic lymphocytes (CTLs) in COVID-19 patients with a reduced ability to produce CD107a, IFN-*γ*, IL-2, granzyme B, and tumour necrosis factor *α* (TNF-*α*).(Zheng et al., 2020) Finally, preliminary unpublished reports described the presence of peripheral blood monocytes with morphologic and phenotypic changes displaying macrophage markers. The severity of the alterations occurring to these atypical monocytes was correlated with patient outcome.(Zhang et al., 2020)

A better knowledge of the immune response against the virus is mandatory, since it has been postulated that severe forms of the disease are associated with a cytokine storm, where monocytes/macrophages play a central role.(Hirano and Murakami, 2020; Giamarellos-Bourboulis et al., 2020; Schulert and Grom, 2015; Mehta et al., 2020) As a consequence, several trials evaluating the efficacy of immunosuppressive or immunomodulatory drugs (tocilizumab, anakinra, sarilumab, eculizumab, ruxolitinib, fingolimod, emapalumab, tofacitinib, meplazumab) are currently undergoing worldwide.(Search of: immune therapy | COVID - List Results - ClinicalTrials.gov)

In attempt to provide our contribution to the field, we designed the present study describing PBMC’s characteristics and basic inflammatory markers values in a cohort of Italian patients who were diagnosed of SARS-CoV-2 infection and were hospitalized.

## Materials and methods

### Study population

We enrolled patients consecutively admitted to the IRCCS Ca’ Granda Ospedale Maggiore Policlinico Foundation, a University Hospital in Milano, Italy, diagnosed with COVID-19 in the period from March 17, 2020, to April 3, 2020. Diagnosis of COVID-19 was defined by as the presence of compatible signs/symptoms (fever, cough, coryza, myalgia, diarrhoea, dyspnoea, tachypnoea, ageusia, anosmia) and a positive nasopharyngeal swab for SARS-CoV-2 detected through real-time reverse transcription-polymerase chain reaction (rRT-PCR). Serum levels of inflammatory biomarkers were assessed as standard clinical practice. Flow cytometry analysis was performed on fresh blood left-over of samples collected at hospital admission for clinical purposes. We analysed only blood samples collected in the first 48 hours since diagnosis. Samples of fresh blood left-over from local blood donor bank were also employed as healthy controls. The study was approved by the Ethical Committee Milano Area 2 (#358_2020) and was conducted in accordance with the Helsinki Declaration.

### Inflammatory biomarkers assessment and clinical data collection

Procalcitonin (PCT), ferritin and interleukin-6 (IL-6) were measured with electro-chemiluminescent immunoassays (ECLIA) on a Roche Cobas e801 instrument (Roche Diagnostics, Monza, Italy). C-reactive protein (CRP) was measured with an immunoturbidimetric method and lactate dehydrogenase (LDH) with the International Federation of Clinical Chemistry (IFCC) optimized method on a Roche Cobas c702 instrument (Roche Diagnostics, Monza, Italy).

For each patient, the P/F ratio (partial pressure of oxygen [PaO_2_]/fraction of inspired oxygen [FiO_2_]), was calculated at the admission to the hospital. The 28-days mortality was collected from electronic medical records.

### SARS-CoV-2 detection

Two different methods were used for viral detection. The first one consisted in Seegene Inc reagents (Seoul, Korea), RNA extraction with STARMag Universal Universal Cartridge kit on Nimbus instrument (Hamilton, Agrate Brianza, Italy) and amplification with Allplex® 2019-nCoV assay, while the second employed a GeneFinder® COVID-19 Plus RealAmp Kit (OSANG Healthcare, Anyangcheondong-ro, Dongan-gu, Anyang-si, Gyeonggi-do, Korea) on ELITech InGenius® instrument (Torino, Italy). Both assays identify the virus by multiplex rRT-PCR targeting three viral genes (E, RdRP and N).

### Flow cytometry analysis

Left-over peripheral blood samples were processed within 24 hours from collection for the following evaluations: (i) classical lymphocyte subpopulations count, (ii) CD4+ T-cells polarization, (iii) lymphocytes activation status, (iv) monocyte subpopulations and (v) NK-cell subsets. For this purpose, we employed the monoclonal antibodies (BD Biosciences, San Jose, CA) reported in supplementary table 1. Each antibody was titrated individually and the optimal dilution for a given staining of 100 μL volume of whole blood was determined by comparison with the isotype matched controls. After the cell staining, erythrocytes were lysed with lysis buffer (Pharmlyse, BD Biosciences). Following the washing, samples were acquired using a BD FACSLyric flow cytometer equipped with three lasers: a 405 nm violet laser, a 488 nm blue laser and a 647 nm red laser. For each tube we set a stopping gate criterion of 50,000 events in the lymphocyte gate, forward scatter (FSC) versus side scatter (SSC), and the data were analysed using FACSSuite and FlowJo softwares (BD Biosciences). An automatic standard compensation was applied for each acquisition. Internal quality assurance procedures included BD cytometer setup and tracking beads, according to the manufacturer’s instructions. Supplementary figure 1 shows the gating strategy employed.

### Statistical analysis

Descriptive statistics were performed for all the variables assessed in the study population. The Spearman test and linear regression were used to examine correlations whereas Kruskall-Wallis test and multiple t test were employed to assess differences among groups. A *p* value <0.05 was deemed statistically significant. All the analysis was performed with GraphPad Prism 8 (GraphPad Inc, USA).

## Results

### Demographic and clinical characteristics and biochemical values

Sixty-three patients with confirmed SARS-CoV-2 infection were enrolled, on average 5.3 days (SD 2.9) after symptoms onset. The median P/F ratio at admission was 170 mmHg. Hypoxemia was severe (P/F < 100) in 13 patients (20.6%), moderate (P/F 100-200) in 13 (20.6%) and mild (P/F 200-300) in 23 (36.5%), while the remaining 14 patients had a P/F > 300 mmHg. Overall, the 28-days mortality rate was 15.9% (10/63). Demographic and clinical characteristics and inflammatory biomarker values are shown in Table 1. Overall, all the inflammatory markers considered were above the reference intervals.

**Table 1.** Demographic and clinical characteristics and inflammatory biomarkers levels of 63 patients with COVID-19 at diagnosis.

| Variable | Value | Reference values |  |
| --- | --- | --- | --- |
| Men, n (%) | 48 (76.2) |  |  |
| Age, mean (SD) | 59.1 (13.7) |  |  |
| Days since symptoms appearance, mean (SD) | 5.3 (2.9) |  |  |
| P/F ratio | 170 (107-290) | >300 | * |
| CRP (mg/dL) | 11.95 (7.43-19.02) | <0.5 | * |
| Ferritin (µg/L) | 1,503 (724-2,887) | 30-400 | * |
| LDH (U/L) | 335 (273-424) | 135-225 | * |
| IL-6 (ng/L) | 81.95 (28.9-112.8) | 0-10 | * |
| PCT (µg/L) | 0.33 (0.16-1.46) | 0.02-0.06 | * |
| Fibrinogen (mg/dL) | 571 (470-722) | 165-300 | * |
| D-dimer (µg/L) | 1,479 (678-2,552) | <500 | * |
| ALT (U/L) | 41 (28-69) | 9-59 |  |

All data are reported as median with interquartile range (IQR) except when stated otherwise.
\* Values outside of normality range. (P/F ratio: partial pressure of oxygen [PaO<sub>2</sub>]/fraction of inspired oxygen [FiO<sub>2</sub>]; CRP: C-reactive protein; LDH: lactate dehydrogenase; IL-6: interleukin 6; PCT: procalcitonin; ALT: alanine aminotransferase)

### White blood cells total count and subpopulations

At diagnosis of SARS-CoV-2 infection, white blood cells (WBC) count, monocytes and neutrophils median values were within the reference intervals employed in our Centre. Instead, lymphocyte counts were well below the lower reference limit. Overall, lymphocytopenia was found in 51 of the 63 patients (80.9%). Consequently, the NLR was oriented toward high values, well above the reference intervals of 0.78-3.53 (Table 2).(Forget et al., 2017)

**Table 2.** White blood cell count, leukocytes subpopulations and reference values employed in our centre.

| Variable | Value | Reference values |  |
| --- | --- | --- | --- |
| **White blood cells (cells/μL)** | 6,520 (5,015-9,200) | 4,800-10,800 |  |
| **Monocytes (cells/μL)** | 330 (195-510) | 300-600 |  |
| **Lymphocytes (cells/μL)** | 720 (520-1,135) | 1200-3,400 | * |
| **Neutrophils (cells/μL)** | 5,070 (3,785-8,105) | 1,500-6,500 |  |
| **Neutrophils/Lymphocytes ratio** | 7.5 (4.39-14.02) | 0.78-3.53 | * |
| **CD3+ lymphocytes (cells/μL)** | 500 (273-728) | 700-2,100 | * |
| **CD4+ lymphocytes (cells/μL)** | 342 (192-451) | 300-1,400 |  |
| **CD8+ lymphocytes (cells/μL)** | 128 (55-215) | 200-900 | * |
| **CD4+/CD8+ ratio** | 2.93 (1.9-3.7) | 1-3.6 |  |
| **CD19+ lymphocytes (cells/μL)** | 120 (54-182) | 100-500 |  |
| **Natural killer cells (cells/μL)** | 67 (35-182) | 90-600 | * |
| **Natural killer T cells (cells/μL)** | 31 (11-73) | 143 (median value) | * |
| **Platelets ($10^3$ cells/μL)** | 247 (162-318) | 130-400 |  |

All data are reported as median with interquartile range (IQR). \* Values outside of reference intervals.

### Lymphocyte subpopulations

Lymphocyte subpopulations mirrored the general feature of lymphocytopenia. Median value of CD3+ and CD8+ cytotoxic lymphocytes (CTLs) were below the lower reference limit and also CD4+ lymphocytes were reduced, but still in the lower part of the reference interval like CD19+ lymphocytes (Table 2). Likewise, the two groups of cytotoxic lymphocytes belonging to the innate immunity, NK and NKT cells, appeared reduced in number and below the lower reference limits (Figure 1 and Table 2). Regarding the activation status of T lymphocytes, assessed through the expression of CD38 and HLA-DR, we observed median frequencies of CD38+HLA-DR+ T helper cells (1.39% of the CD4+ cells, IQR 0.7-2.89) in the lower part of the reference interval (1.3-1.9%). Instead, the median frequencies of CD38+HLA-DR+ CTLs (6.75% of the CD8+ cells, IQR 3.92-10.07) were above the upper reference limit (1.45-5.7%).

**Figure 1.**
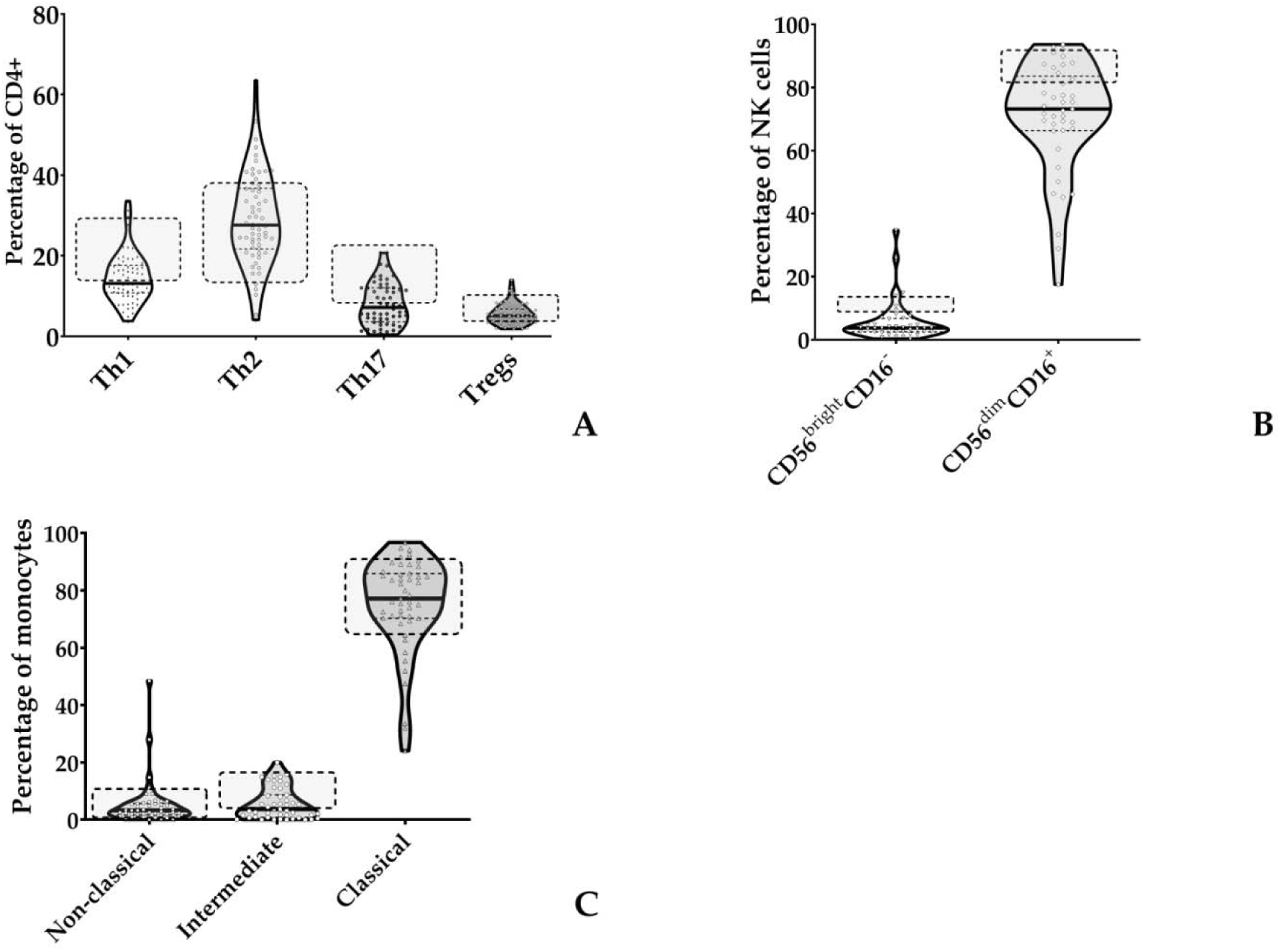
Violin plots showing lymphocyte and monocyte subpopulations (reported as median value and IQR) assessed in 63 patients with COVID-19 at diagnosis. Reference values are represented as dotted regions: (**A**) CD4+ cells differentiation; (**B**) NK cell subpopulations; (**C**) monocytes subpopulations. (IQR: interquartile range; Th1: Type 1 T helper cells; Th2: Type 2 T helper cells; Th17: T helper 17 cells; Tregs: regulatory T cells; NK: natural killer cells).

The T CD4+ lymphocyte subclasses are presented in Figure 1. The relative median frequencies of Th1, Th2, Th17 and Treg lymphocytes were respectively 13.12% (IQR 10.80-17.60), 27.6% (IQR 21.67-36.70), 7.2% (IQR 3.59-12) and 5.1% (IQR 3.61-6.8), respectively. Reference values for the same cell populations are 15-28%, 12-38%, 9-24%, 5-11% of CD4+ cells for Th1, Th2, Th17 and Treg lymphocytes, respectively. Consequently, the median Th1/Th2 ratio was altered whereas the Treg/Th17 ratio was not, being 0.47 (IQR 0.36-0.65; reference values 0.7-1.2) and 0.62 (IQR 0.47-1.21; reference values 0.3-1.7), respectively.

### Natural killer cells

As stated above, NK cells median values were reduced and below the lower reference value. The frequencies of NK cell subsets are depicted in Figure 1 B. The median NK CD56^bright^CD16-(immature) and NK CD56^dim^CD16+ (mature) frequencies were 3.7% (IQR 2.5-6.75) and 73.2% (IQR 66.3-83.6). Within immature NK cells, 30% of patients showed more than 1% of terminally differentiation marker (CD57) (IQR 0-2.9), whereas in mature NK cells CD57 was expressed on 40% of these cells (IQR 22.25-54.65). Reference values are considered a median of 10% for NK CD56^bright^CD16- and a range comprised between 80-90% for NK CD56^dim^CD16+. A range of 30-60% of NK CD56^dim^CD16+ usually express CD57.(Lopez-Vergès et al., 2010)

### Monocytes

The median frequencies of monocyte subpopulations are displayed in figure 1 C. Overall, relative subpopulations were 3.3% (IQR 1.66-5.7), 3.7% (IQR 0.8-8.7) and 77.2% (IQR 70.28-85.9) for non-classical monocytes (CD14^dim^CD16+), intermediate monocytes (CD14+CD16+) and classical monocytes (CD14+CD16-), respectively. Reference intervals for the above monocyte populations ranging between 1-10.2%, 4.7-17.8% and 69.8-92.8% for non-classical, intermediate and classical monocytes, respectively. Finally, the median prevalence of monocytes HLA-DR+ was 83.4% (IQR 75.2-92.3), below the usual median value of 98%.

Morphological identification of monocytes in COVID-19 patients became challenging due to altered scatter properties. Hence, we modified the monocyte identification strategy including a gate for CD4^dim^ cells (Supplementary Figure 1, panel E). Furthermore, we noticed distinct CD4^dim^ cell population with high FSC and SSC (Figure 2 A). Blood smear examination confirmed the presence of atypical monocytes with vacuoles in their cytoplasm (Figure 2 B). Therefore, we modified flow cytometer parameters with reduced photomultiplier tube values in order to analyse better these peculiar monocytes (high-SC). We performed this analysis in a subgroup of 14 patients and we considered significant a value of high-SC > 2% of total monocytes. Overall, the high-SC population was present in 11/14 (79%) patients where they represented a variable fraction of total monocytes (range 2-14%).

**Figure 2.**
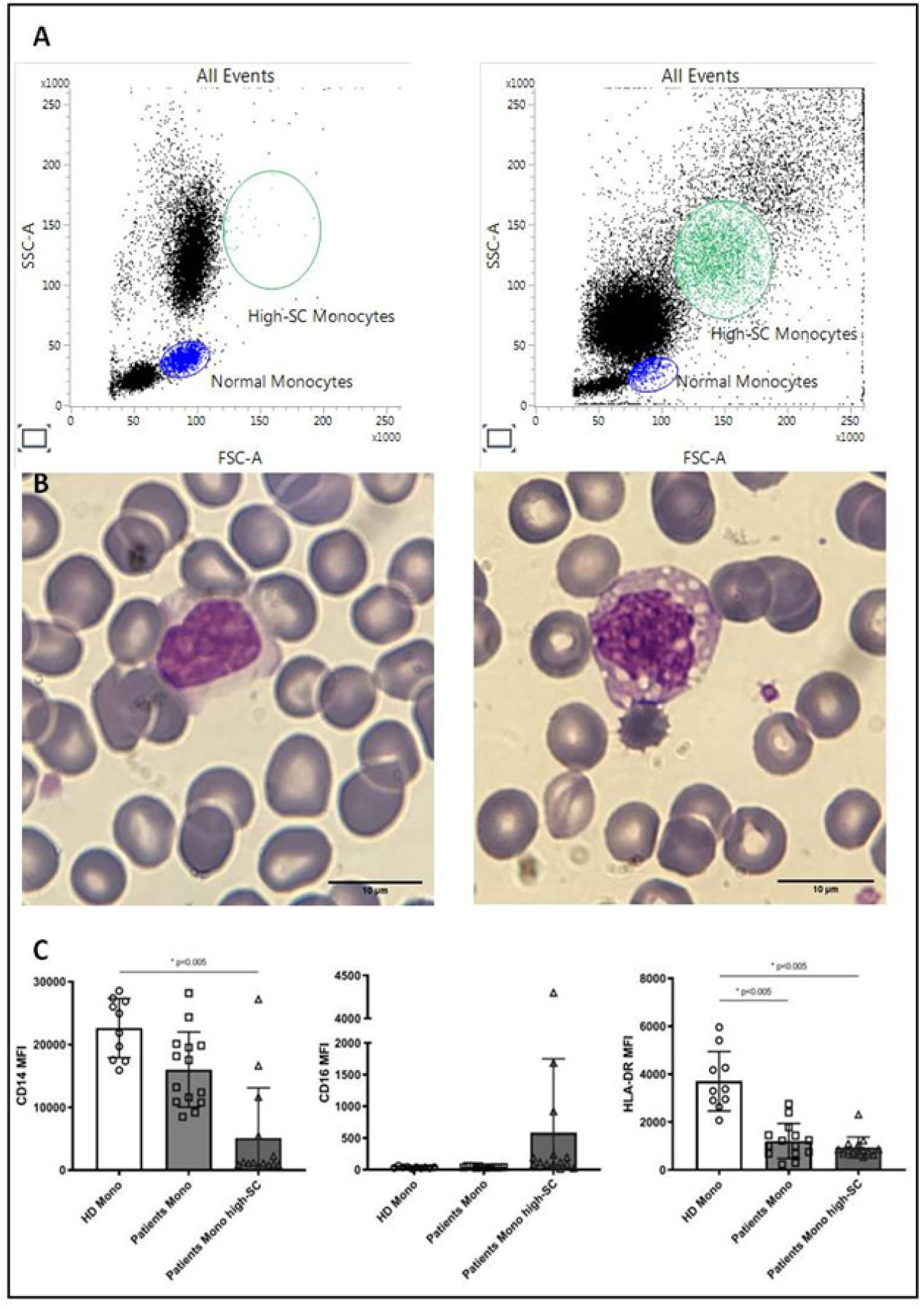
Morphologic and phenotypic differences between monocytes from a healthy donor (left) and a COVID-19 patient (right): **(A)** flow cytometry dot plot showing normal and high-SC monocytes; **(B)** monocytes from May-Grunwald Giemsa stained blood smears (100X magnification, Leica Microscope DMLS); **(C)** histograms showing median fluorescence intensity (MFI) of CD14, CD16 and HLA-DR on monocytes from healthy donors (HD, n=10), normal and atypical monocytes from COVID-19 patients (n=14). (* *p* value <0.005)

To better understand the differences in the phenotypic signature between normal and high-SC monocytes in COVID-19 patients and monocytes from healthy donors (n=10), we compared the Median Fluorescence Intensity (MFI), a parameter proportional to antigen density, of CD14, CD16 and HLA-DR in these monocyte populations. Overall, the MFI of CD14 and HLA-DR on high-SC monocytes was lower than MFI on monocytes of healthy donors (*p*<0.0001). Regarding HLA-DR, we also found a reduction on monocytes with normal scatter properties from COVID-19 patients in comparison to monocytes of healthy donors (*p*=0.0008). Finally, for CD16 we observed a rising trend of MFI in high-SC monocytes compared to normal monocytes from COVID-19 patients and healthy donors (Figure 2 C).

Correlations with inflammatory biomarkers

We also assessed the relationship between the different variables evaluated in our cell populations and inflammatory biomarkers. Interestingly, the serum values of IL-6 and the median monocytes count were not correlated (R=0.01; *p*=0.60). Instead, the serum values of IL-6 showed a moderate correlation with the median frequency of Th2 lymphocytes (R=0.17; *p*=0.04). (Supplementary Figure 3) All other variables did not show any correlation.

### Variables associated with death

Finally, we stratified all the results presented above based on 28-days mortality. Patients who died within 28 days from admission had higher age, LDH values, CD4/CD8 ratio and percentages of CD38+/HLA-DR+ CTL compared to those alive (Table 3). Instead, they had lower P/F ratio, CD3+ and CD4+ lymphocyte counts.

**Table 3.** Variables with significant differences between those who died in the 28 days from admission and those who did not.

|  | *p* value | Mean<br>dead<br>(n=10) | Mean<br>alive<br>(n=53) | Difference | SE of<br>difference | t ratio | df |
| --- | --- | --- | --- | --- | --- | --- | --- |
| **Age (years)** | 0.002 | 71.3 | 56.8 | 14.5 | 4.4 | 3.3 | 61.0 |
| **LDH (U/L)** | 0.024 | 484.2 | 338.2 | 145.9 | 61.2 | 2.4 | 28.0 |
| **P/F admission** | 0.043 | 138.1 | 204.6 | -66.5 | 32.2 | 2.1 | 61.0 |
| **CD3+ (cells/μL)** | 0.028 | 337.4 | 585.9 | -248.6 | 110.5 | 2.2 | 57.0 |
| **CD4+ (cells/μL)** | 0.042 | 232.3 | 381.1 | -148.8 | 71.6 | 2.1 | 57.0 |
| **CD4/CD8 ratio** | 0.015 | 5.4 | 3.0 | 2.4 | 0.9 | 2.5 | 57.0 |
| **CD38+/HLA-**<br>**DR+ on CD8+**<br>**cells (%)** | 0.026 | 13.5 | 7.6 | 5.8 | 2.5 | 2.3 | 60.0 |

(LDH: lactate dehydrogenase; P/F: partial pressure of oxygen [PaO<sub>2</sub>]/fraction of inspired oxygen [FiO<sub>2</sub>])

## Discussion

In our cohort of patients hospitalized for COVID-19, we observed a reduction of circulating lymphocytes, both in terms of total count and specific subpopulations. Precisely, all cytotoxic lymphocytes (CTLs, NK cells and NKT cells) were below the lower reference limits. Moreover, the number of activated CD8+ T cells was slightly increased, whereas the T CD4+ cell population was polarized toward a Th2 phenotype. Intriguingly, monocytes displayed morphological and phenotypical alterations. In fact, they had higher scatter properties, and a reduced percentage of immunocompetent cells compared to the healthy donors. IL-6 levels were not correlated to monocyte count but instead to T cells with a Th2 phenotype. Finally, patients who died within 28 days from admission presented lower counts of CD3+ and CD4+ cells and higher percentages of activated T cells when compared to those alive.

Our results are in line with those recently reported by several groups regarding a profound reduction of lymphocytes in the context of a preserved total WBC count.(Chen et al., 2020; Giamarellos-Bourboulis et al., 2020; Zhou et al., 2020) Of note, in our cohort lymphocytopenia was present in 51 out of 63 of the enrolled patients (81%), which is a higher percentage compared to those observed in the works by Chen (72% of severe cases, 10% of moderate cases) and by Huang (63% of all cases).(Chen et al., 2020; Huang et al., 2020) The reduction of cytotoxic lymphocytes, both of innate and adaptive immunity, is a relevant finding. Indeed, several models of macrophage activation syndrome in the context of rheumatologic diseases highlighted how cytolytic cells may induce apoptosis in activated macrophages and T cells and how cytolysis serves to control the inflammatory response.(Crayne et al., 2019) An impairment in the cytolytic function may result in a overstimulation of the immune system leading to the multiorgan failure and this defect has been linked to elevated levels of IL-6.(Cifaldi et al., 2015)

Regarding monocytes, Zhang and Zhou reported in their studies (Zhang et al., 2020; Zhou et al., 2020) a relative increase in intermediate and non-classical monocytes. Instead, we detected normal value of classical and non-classical monocytes and a relative reduction of intermediate monocytes. It should be underlined that our analysis was performed in samples collected early in the course of the disease, while it is unclear when the samples were collected in the studies conducted in China. Similarly to these studies, the monocyte features observed in our cohort showed not only a shift in the forward scatter but also in the side scatter properties, a parameter related to cell complexity. These high-SC monocytes displayed a decrease of CD14 and HLA-DR antigen density. Notably, we also detected a reduced expression of HLA-DR, either as percentage or fluorescence intensity, on patients’ monocytes with normal scatter properties. The same observation was made by Giamarellos-Bourboulis and colleagues, which also demonstrated an inverse correlation between HLA-DR molecules on CD14-monocytes of patients with COVID-19 and serum levels of IL-6.(Giamarellos-Bourboulis et al., 2020) Of note, values lower than 30% of HLA-DR+ monocytes have become an accepted definition of immune-paralysis in the context of sepsis.(Frazier and Hall, 2008) It will be fascinating to follow this population of atypical monocytes during the course of the disease.

Regarding lymphocytes, Zhou and colleagues observed that CD4+ T lymphocytes were rapidly activated to become pathogenic Th1 cells, secreting proinflammatory cytokines. The resulting environment induced inflammatory CD14+CD16+ monocytes with high expression of IL-6.(Zhou et al., 2020) In contrast, in our study the T helper ratio was oriented toward a Th2 polarization and Il-6 levels were directly proportional to this population. This is an expected finding, considering that IL-6 shift the Th1/Th2 balance of an immune response towards Th2, by promoting early IL-4 expression and by rendering CD4 T cells unresponsive to IFNγ signals.(Dienz and Rincon, 2009) Likewise, Giamarellos-Bourboulis et al. did not detect an immune response oriented toward a Th1 phenotype in their cohort, with IFNγ values below the detection limit in all the patients.(Giamarellos-Bourboulis et al., 2020)

Finally, we observed some peculiar results stratifying the variables according to 28-days mortality. Indeed, in addition to the clinical variables (age, P/F ratio, LDH) which are expected to be linked with the disease outcome, we have observed how lymphocytopenia and lymphocyte activation status are more evident in those patients who died. Taken together those data highlight that a worst outcome is associated with a depleted lymphocytes compartment, partially activated and thus unable to eliminate/abrogate an inflammatory stimulus (i.e activated monocytes).

It should be noted that samples for this study were collected at diagnosis, on average after 5 days from the appearance of symptoms. Considering that the mean incubation time of COVID-19 after infection has been estimated to be 5 days, we can assume that on average our patients were in the 10th day of infection. For this reason, our results could be considered a faithful description of the immunological changes occurring in the early phases of the disease and could complement well the data provided by others. At the same time, we acknowledge that our research has limitations. First, it is a single centre study, conducted on a small number of patients. Second, it involved only patients diagnosed in hospital, therefore with a severe disease. Third, we did not investigate the presence and the features of immune cells at alveolar level, where most of COVID-19’s immunopathology occurs. To overcome this problem, we are collecting and analysing both PBMC and bronchoalveolar lavage fluid samples of COVID-19 patients.

Overall, based on our results, we can hypothesize that COVID-19 causes a reduction of the lymphocyte populations, especially the cytotoxic lymphocytes, thus affecting both the innate and adaptive immunity. The diminished number of CTLs, supported by the elevated levels of cytokines such as IL-6, is probably associated with a decrease in global cytotoxic activity, which in turn can lead to a failure in the removal of the triggering antigens and therefore can elicit an overstimulation of the immune system. Monocytes, considered the crucial players in the pathogenesis of lung damage caused by the cytokines storm, showed substantial phenotypical alterations already in the early phase of the disease. The mechanism leading to monocytes alteration is unclear, but a direct viral role can be suspected. Indeed, Zhang et al.(Zhang et al., 2020) showed how monocytes express ACE2, the entry receptor of SARS-CoV-2, and it has been reported how other viral infections (influenza A virus, vaccinia virus, vesicular stomatitis virus) can trigger rapid and substantial differentiation of monocyte profile toward dendritic cells.(Hou et al., 2012)

In a similar manner, our data might suggest that, in the early phases of COVID-19, the virus can elicit an inflammatory response leading to a reduction in the number of both cytotoxic lymphocytes (CTL, NK cells, NKT cells) and helper lymphocytes. These changes, which are more pronounced in patients with adverse outcome, could represent the first steps in the cascade leading to the uncontrolled activation of monocytes in the context of cytokines storm.

In conclusion, we present here the first description of immunologic features at diagnosis of a cohort of Italian COVID-19 patients that both confirm already available evidence and add novel elements to the COVID-19 jigsaw. The interpretation of our data opens interesting new perspectives for therapeutic interventions. It is possible to speculate that an early initiation of an immunomodulatory treatment might abrogate the immune system alterations leading to monocytes activation-dysfunction and to the consequent lung damage. Nevertheless, further prospective studies are needed to provide a precise picture of the immunologic profile during the course of the disease in order to define better therapeutic approaches and to improve the clinical management of COVID-19 patients.

## Data Availability

Alla data reported are available for further analysis.

## Authors contributions

AL, LP, AB and AG conceived the study. VC, EP, GL, GC, VS, GG, RG and MM enrolled the patients. LP, AC, ET, MT and FC performed the immunologic and biochemical analysis. AL, LP and AB analysed the data. AL and LP wrote the first draft. All authors reviewed the final version of the manuscript.

## Acknowledgments

The authors are thankful to Luca Vecchia for the critical review of the draft and Antonella Montemurro and Francesca Marangoni for technical support.

## “COVID-19 NETWORK” WORKING GROUP

**Fondazione IRCCS Ca’ Granda Ospedale Maggiore Policlinico**. <u>Scientific Direction:</u> Silvano Bosari, Luigia Scudeller, Giuliana Fusetti, Laura Rusconi, Silvia Dell’Orto; <u>Department of Transfusion Medicine and Hematology (Biobank):</u> Daniele Prati, Luca Valenti, Silvia Giovannelli; <u>Infectious Diseases Unit:</u> Andrea Gori, Alessandra Bandera, Antonio Muscatello, Davide Mangioni, Laura Alagna, Giorgio Bozzi, Andrea Lombardi, Riccardo Ungaro, Teresa Itri, Valentina Ferroni, Valeria Pastore, Roberta Massafra, Ilaria Rondolini; <u>Angelo Bianchi Bonomi Hemophilia and Thrombosis Center and Fondazione Luigi Villa:</u> Flora Peyvandi, Roberta Gualtierotti, Barbara Ferrari, Raffaella Rossio, Elisabetta Corona, Nicolo Rampi, Costanza Massimo; <u>UOC Internal Medicine, Immunology and Allergology</u>: Nicola Montano, Barbara Vigone, Chiara Bellocchi, Elisa Fiorelli, Valerie Melli, Eleonora Tobaldini; <u>Respiratory Unit and Cystic Fibrosis Adult Center:</u> Francesco Blasi, Stefano Aliberti, Maura Spotti, Edoardo Simonetta, Leonardo Terranova, Francesco Amati, Carmen Miele, Sofia Misuraca, Alice D’Adda, Silvia Della Fiore, Marta Di Pasquale, Marco Mantero Martina Contarini, Margherita Ori, Letizia Morlacchi, Valeria Rossetti, Andrea Gramegna, Maria Pappalettera, Mirta Cavallini, Annalisa Vigni; <u>Cardiology Unit:</u> Marco Vicenzi, Irena Rota. Emergency Unit: Giorgio Costantino, Monica Solbiati, Ludovico Furlan, Marta Mancarella, Giulia Colombo, Giorgio Colombo, Alice Fanin; <u>Acute Internal Medicine:</u> Valter Monzani, Angelo Rovellini, Laura Barbetta, Filippo Billi, Christian Folli; <u>Rare Diseases Center:</u> Marina Baldini, Irena Motta, Natalia Scaramellini; <u>General Medicine and Metabolic Diseases:</u> Anna Ludovica Fracanzani, Rosa Lombardi, Federica Iuculano; <u>Geriatric Unit:</u> Matteo Cesari, Marco Proietti, Laura Calcaterra. **Istituto di Ricerche Farmacologiche Mario Negri IRCCS**: Alessandro Nobili, Mauro Tettamanti, Igor Monti.

---

**Supplementary Figure 1.**
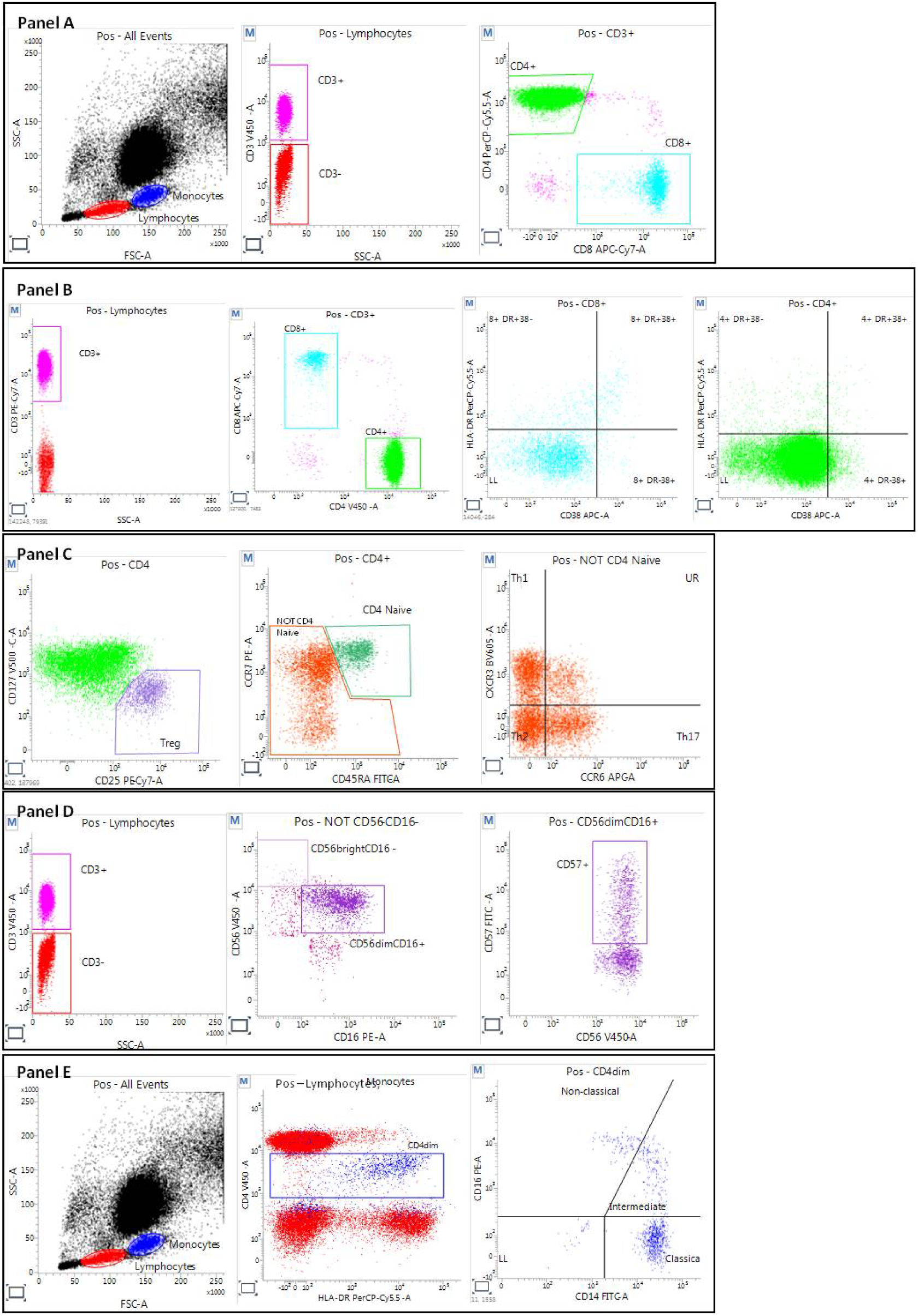
Gating strategy employed. Lymphocytes were firstly gated on the basis of their scatter properties (Side Scatter, SSC and Forward Scatter, FSC) and then divided in CD3+ cells (T lymphocytes) and CD3- cells; subsequently T-helper cells (CD4+) and T cytotoxic cells (CD8+) were gated in T lymphocyte population (panel A); in both CD4+ and CD8+, we also evaluated the expression of the activation markers CD38 and HLA-DR (panel B).

Within the CD4+ cell population we identified T regulatory lymphocytes (Treg, CD25++CD127low) and, after the exclusion of CD4+ naïve T cell (CD4+CCR7+CD45RA+), as suggested by EQAs for Cellular Immunodeficiency Diagnostic Program (INSTAND), we evaluated the distribution of TH1 (CXCR3+CCR6-), TH2 (CXCR3-CCR6-) and TH17 (CXCR3-CCR6+) cell populations (panel C).

Within CD3- cells and after the exclusion of CD56/CD16 double negative cells, we divided NK cells according to the expression of CD56 and CD16 in immature NK cells (CD56^bright^CD16-) and mature NK cells (CD56^dim^CD16+). We then evaluated the expression of CD57 on this latter population to determine the terminally differentiated NK cells (panel D).

For the analysis of monocyte populations, after the classical gate on morphological properties, we identified cells based on dim expression of CD4. Monocytes were then evaluated for the expression of HLA-DR (immunocompetent monocytes), and for CD14 and CD16 and divided in classical (CD14+CD16-), intermediate (CD14+CD16+) and non-classical monocytes (CD14^dim^CD16+) (panel E).

**Supplementary Figure 2.**
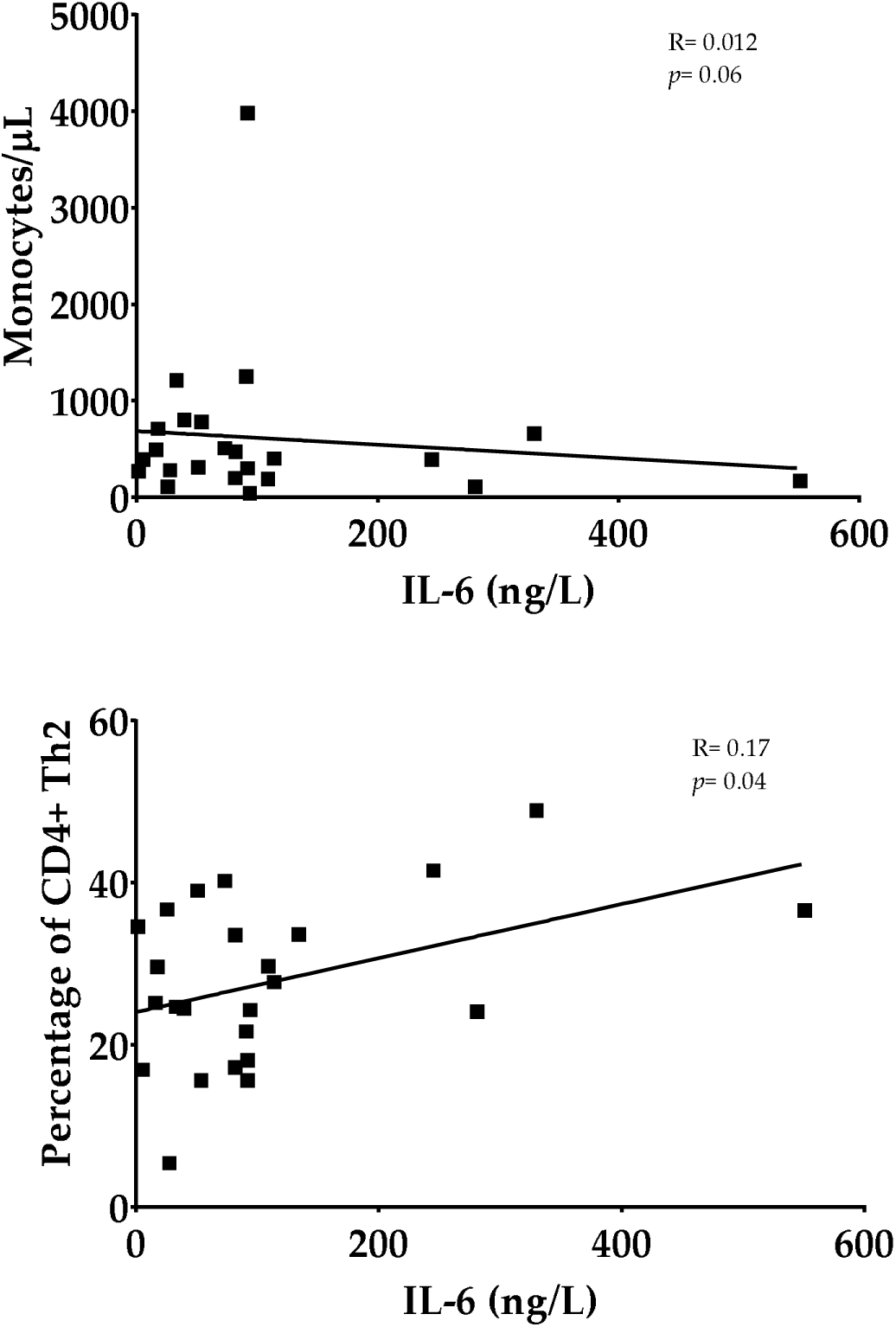
In a subgroup of 24 patients we evaluated the correlation between IL-6 serum values and WBC populations. The IL-6 serum values were unrelated to the monocytes count. Instead, a direct correlation existed between the IL-6 serum values and the prevalence of CD4+ cells with Th2 polarization. (R= linear regression; *p= p* value).

**Supplementary Table 1.** List of the employed antibodies.

|  | **Antigen** | **Clone** | **Fluorochrome** |
| --- | --- | --- | --- |
| **i) Multitest 6-color TBNK commercial Kit** |  |  |  |
| **ii) CD4+ T-cells polarization** | CD45RA | L48 | FITC |
|  | CCR7 | 150503 | PE |
|  | CD4 | SK3 | PerCP-Cy5.5 |
|  | CD25 | M-A251 | PE-Cy7 |
|  | CCR6 | 11A9 | AlexaFluor647 |
|  | CD8 | SK1 | APC-Cy7 |
|  | CD3 | UCHT1 | V450 |
|  | CD127 | HIL-7R-M21 | BV510 |
|  | CXCR3 | 1C6/CXCR3 | BV605 |
| **iii-iv) T Lymphocyte Activation and Monocytes** | CD14 | MΦP9 | FITC |
|  | CD16 | B73.1 | PE |
|  | HLA-DR | L243 | PerCP-Cy5.5 |
|  | CD3 | SK7 | PE-Cy7 |
|  | CD38 | HB-7 | APC |
|  | CD8 | SK1 | APC-Cy7 |
|  | CD4 | SK3 | V450 |
| **v) NK** | CD57 | HNK-1 | FITC |
|  | CD16 | B73.1 | PE |
|  | CD3 | SK7 | PE-Cy7 |
|  | CD56 | B159 | V450 |

## Notes

### Competing Interest Statement

The authors have declared no competing interest.

### Funding Statement

The grant “COVID-19 Biobank” was provided by the Scientific Direction, Foundation IRCCS Ca' Granda Ospedale Maggiore Policlinico, Milano, Italy.

